# Prognostic implications of Global Longitudinal Strain versus Ejection Fraction in End-stage Renal Disease: a systematic review protocol

**DOI:** 10.1101/2022.07.29.22278210

**Authors:** Barbara Leticia da Silva Guedes de Moura, Anderson Gusthavo dos Santos Mucenieks, Enrico Prajiante Bertolino, Giordanna Chiqueto Duarte, João Vitor Dudek, Mariana Amâncio Daniel da Silva, Izabel Galhardo Demarchi, Sergio Seiji Yamada, Rogerio Toshiro Passos Okawa, Jorge Juarez Vieira Teixeira

## Abstract

**Background:** In hemodialysis (HD) patients, the presence of Heart failure (HF) at the start of dialysis is a strong and independent predictor of short and long-term mortality, and its prevalence increases with declining kidney function and HD time. Left ventricular (LV) ejection fraction (EF) is widely used as a measure of systolic function. Reduced EF (<50%) is an important prognostic marker, however, less than 15% of End-stage Renal Disease (ESRD) patients have detectable systolic dysfunction and the EF is susceptible to loading conditions, which change dramatically during interdialytic intervals. Global Longitudinal Strain (GLS) derived by 2D Speckle-Tracking Echocardiography (STE) is an emerging technique for measuring more subtle disturbances in LV systolic function. Although ESRD patients have subclinical evidence of impaired strain but preserved EF, there is evidence that GLS is better in ESRD group receiving maintenance HD compared with moderate-advanced CKD patients. This systematic review will evaluate the evidence related to the incremental prognostic value of LV GLS by 2D-STE concerning mortality and cardiovascular (CV) events in ESRD patients.

**Methods:** This protocol is reported according to the PRISMA-P guideline. The databases PubMed, EMBASE, LILACS, Web of Science, and Google Scholar system will be searched and double screening for longitudinal studies that assessed the prospective association of STE-derived parameters with at least one of the pre-specified outcomes in ESRD patients. Discrepancies will be resolved through consensus. A modified version of the Newcastle-Ottawa Quality Assessment Scale of cohort studies will be used. We intend to use the random-effects model, considering at least moderate heterogeneity between studies. If data allow, we will perform meta-regression to explore potential sources of between-study heterogeneity. An adaptation of the GRADE framework for prognostic studies will be employed to judge the quality of evidence for each outcome reported in this systematic review.

**Discussion:** This systematic review will summarize current evidence about STE-derived measures in the ESRD patients and clarify the incremental prognostic value of this diagnostic tool versus LVEF in these patients. Evidence about other measures (circumferential and radial strain) or 3D STE-derived indices will also be investigated.

**Systematic review registration:** This systematic review protocol was submitted to the PROSPERO international registry of systematic reviews (Registration number: CRD42020222505).

**Strengths and limitations of this study**

→ The PROSPERO registry helps to promote and maintain transparency in the process and to assist in minimizing the risk of bias. Four databases and one additional grey literature source helps ensure more complete coverage of the topic. A modified version of the Newcastle-Ottawa Quality Assessment Scale of cohort studies will be used and an adaptation of the GRADE framework for prognostic studies to assess the quality of evidence for each outcome reported will be applied. All the review phases will be peer-reviewed and will be validated by methodological and clinical experts.
→ Left ventricular (LV) ejection fraction (EF) is widely used as a measure of systolic function. Reduced EF (50%) is an important prognostic marker, however, less than 15% of End-Stage Renal Disease (ESRD) patients have detectable systolic dysfunction. Global Longitudinal Strain (GLS) derived by 2D Speckle-Tracking Echocardiography (STE) is an emerging technique for measuring more subtle disturbances in LV systolic function, however, its incremental prognostic value to ESRD is controversial.
→ This study has limitations. Because heterogeneity is quite common in these systematic reviews, there may be limited scope for meta-analysis. Possibly, the spectrum of patients in different research sites is the main factor, in most cases, but heterogeneity in associations between studies may be difficult to explain.
→ This systematic review will evaluate the evidence related to the GLS incremental prognostic value in End-Stage Renal Disease patients, which may lower morbidity and mortality and influence its use as a prognostic marker for ESRD patients.

## Background

Cardiovascular disease (CVD) is the leading cause of morbidity and mortality in patients with advanced chronic kidney disease (CKD), accounting for >50% of all deaths.[1] Although classical Framingham risk factors remain important, there is increasing evidence to support renal-specific and dialytic disturbances increases the cardiovascular risk in this population, such as sodium/fluid balance, hyperuricemia, inflammation, vascular calcification, and abnormal bone mineral metabolism [2], and this complex interaction results in a high prevalence of abnormalities of cardiac structure and function in patients with End-stage Renal Disease (ESRD) [3].

Though hemodialysis (HD) should theoretically improve cardiovascular function by correcting fluid overload and small molecule accumulation, cardiovascular mortality continues to be disproportionately high in the HD population [4]. The impact of renal replacement therapy (RRT) on cardiovascular function and injury is not well understood and may inadvertently be contributing to the accelerated development of type 4 cardiorenal syndrome (CRS): CKD leading to an impairment of cardiac function. The unique physiology of cardiovascular abnormalities in dialysis patients remains poorly understood and several more recently recognized factors, including altered lipid metabolism and accumulation of gut microbiota-derived uremic toxins, also affect cardiovascular function in the context of renal failure [5].

Left ventricular (LV) ejection fraction (EF) is widely used as a measure of systolic function. Reduced EF (<50%) is an important prognostic marker, however, less than 15% of ESRD patients have detectable systolic dysfunction [6] and the prognostic ability of EF can be limited because it is susceptible to loading conditions [7], which change dramatically during interdialytic intervals in hemodialysis patients [8].

Global Longitudinal Strain (GLS) derived by Two-dimensional (2D) Speckle-Tracking Echocardiography (STE) is an emerging technique for measuring more subtle disturbances in LV systolic function. There is emerging data that GLS is a superior predictor of cardiac events and all-cause mortality compared with EF in the general population with and without heart failure (HF) [9,10] even among HF patients with preserved LVEF [11]. Impaired GLS was proved to be associated with microvascular ischemia, interstitial fibrosis, and cardiac myocyte hypertrophy caused by urotoxin, hypertension, and hemodialysis-related myocardial stunning [12-14] which would all lead to cardiovascular mortality and morbidity.

Although studies have demonstrated that patients with moderate-to-severe CKD and dialysis patients have subclinical evidence of impaired strain but preserved EF [14,15] there is evidence that GLS is better in ESRD group receiving maintenance HD compared with moderate-advanced CKD patients [16]. Dialysis patients are susceptible to dramatic changes in loading conditions during interdialytic intervals [8]. Loading conditions may be affect not only EF but also strain values [17] and although GLS independently predicted the outcome in dialysis patients, it did not show incremental prognostic value over the EF on the model with AROii score (a validated 2-year mortality risk score in the hemodialysis cohort) [18].

The prognostic value of LV-GLS in the general CKD patients was demonstrated in recent a meta-analysis [19], but considering the insufficient data until the research date, they did not further analyze GLS in different stages of CKD, especially in ESRD patients.

As the mechanism and applications of abnormal GLS in HD patients have not been fully clarified, we will perform a systematic review of LV GLS as a predictor of total and cardiovascular mortality and morbidity in the ESRD population specifically.

## Methods/Design

This protocol is reported according to the Preferred Reporting Items for Systematic Reviews and Meta-Analyses Protocols (PRISMA-P) [20] and the corresponding checklist used (Supplementary file 1). This includes details of inclusion and exclusion criteria, a data extraction tool, and analytical methods. This systematic review protocol was submitted to the PROSPERO international registry of systematic reviews (Registration number: CRD42020222505).

### Search strategy

The databases PubMed, EMBASE, LILACS (Latin American and Caribbean Literature in Health Sciences), Web of Science, and Google Scholar system (grey literature) and reference lists of eligible studies will be searched for publications until the search date. The Search strategy were determined independently by three authors (BLSGM, RTPO, and JJVT) and were developed first for the PubMed database and the effectiveness will be tested and refined accordingly. The search strategy was developed using two combined blocks of keywords/subject headings (MeSH terms/PubMed) with the Boolean operators OR/AND: (a) Block 1 – “global longitudinal strain” OR “speckle tracking echocardiography”, (b) Block 2 – “chronic kidney disease” OR [‘haemodialysis’ OR ‘renal dialysis’ OR ‘renal AND dialysis’ OR ‘renal dialysis’ OR ‘hemodialysis’] OR “end-stage renal disease”], (c) (a) AND (b). A detailed draft for the search strategy to be used is detailed in Supplementary file 2. The search will be limited to original research reports in peer-reviewed journals and only full-texts, with no language and publication date restriction, will be retained.

### Eligibility criteria

All longitudinal studies that assessed the prospective association of STE-derived parameters with: (a) at least one of the pre-specified outcomes in ESRD (>18 years) population, (b) following ethical standards, will be included and (c) Abstracts, reviews, news, editorials, report cases, conference proceedings or letters to the editor will be excluded.

Search results from each database will be combined and duplicates will be removed before the screening. References will be aggregated into a reference manager tool (EndNote Web) [21]. Additional papers will be identified by searching the reference lists of relevant articles and their citation metrics.

### Outcomes and Measures of effect

The primary outcome will be all-cause mortality whereas secondary outcomes were (1) composite cardiovascular or (2) cardiac endpoints, defined as:

(1) composite CV endpoints, including any combination of CV mortality, coronary heart disease (CHD) events (myocardial infarction-MI, unstable angina, angina/ischemia requiring emergent hospitalization or revascularization), HF hospitalization, new-onset atrial fibrillation (AF), life-threatening arrhythmia, recorded automatic implantable cardioverter-defibrillator (AICD) shocks, stroke, transit ischemic attack or peripheral arterial disease with arterial revascularization procedure;

(2)composite cardiac endpoints, including any combination of CV mortality, CHD events (MI, unstable angina, angina/ischemia requiring emergent hospitalization or revascularization), HF hospitalization, new-onset AF, life-threatening arrhythmia, recorded AICD shocks.

Tertiary outcomes will be any individual secondary endpoint not included in the composite cardiac or CV endpoint.

Due to the characteristics of Doppler and CMR (Cardiovascular magnetic resonance imaging) -based strain, we only will be included studies that calculated strain by STE. A draft for the Eligibility and Data Extraction Form to be used is detailed in Supplementary file 3.

We will provide a narrative synthesis of the findings from the included studies. We will provide summaries of intervention effects for each study by calculating risk ratios (for dichotomous outcomes) or standardized mean differences (for continuous outcomes) from the data presented in the published studies or obtained from study authors.

### Study reviews and appraisals

Initial title and abstract screening will be performed using Rayyan software [22] and full texts of selected articles will be retrieved and double screened for eligibility and data extraction by six researchers working independently (AGSM, BLSGM, EPB, GCD, JVD, MADS). Discrepancies will be resolved through consensus or the opinion of a third review author (JJVT or RTPO) will be sought. For multiple study publications from the same patient cohort, we will choose the study with the largest number of cases, or which one that reported the results separately according to the CKD stage, once there are many studies whose results of patients with early-to-moderate CKD stage are reported together with ESRD patients, which makes it impossible to correctly analyze these subgroups. For studies that presented different outcomes, we will be extracted outcomes from both publications.

Reasons for study exclusion will be documented. Data extraction will be validated by methodological and clinical experts (IGD, JJVT and RTPO, SSY, respectively). Also, information reported in the grey literature will be sought. We will report our search terms as an appendix to published studies.

### Data extraction

The following data will be extracted: (1) citation details (title, type, and year of publication); (2) study details (name, region, and design of the study, sample size); (3) participant details including demographics (age, sex, ethnicity, and follow-up duration), clinical (hypertension, diabetes mellitus-DM, dyslipidemia, smoking status, and known CVD), and laboratory variables; (4) exposure details (hardware and the software used for the acquisition and analysis as well as details regarding the measured LV strain); (5) details of outcomes, including how these were ascertained; (6) statistical methods used, as well as statistics related to the association of interest, and (7) information related to adjustment for potential confounders.

If samples are reported at multiple time points within a study, then each one will be considered individually. If these data are not available, we will extract the association measures that were used. If study data, critical to our proposed analysis, is unclear or absent, the relevant authors will be contacted for clarification.

### Quality assessment, data synthesis, and exploration of heterogeneity

A modified version of the NOS (Newcastle-Ottawa Quality Assessment Scale of cohort studies) will be used to assess the quality of included papers (Supplementary file 4) [23]. Statistical analyses will be performed using Review Manager 5.4 software. This information will be summarized graphically in the final review document. Studies will not be excluded from the main meta-analysis based on the risk of bias.

The degree of heterogeneity will be assessed using the Higgins Thompson I^2^ test [24] and Cochran’s Q test [25]. If possible, according to the included studies, a quantitative synthesis using random-effect meta-analysis will be used to pool the results of the association of interest. For presenting the results, we will likely use the HR (95%CI) as an effect estimate, and will present the data graphically in forest plots. If data allow, we will perform meta-regression to explore potential sources of between-study heterogeneity: (1) study quality, (2) variation in exposure measurement (a type of strain-longitudinal, circumferential or radial strain), and (3) variation in a CV and cardiac outcomes definition. Possible publication bias will be assessed using a Funnel plot [26]. We will assess evidence of publication bias using Egger’s weighted regression method for continuous outcomes, and Begg’s rank correlation test for dichotomous outcomes. Statistical analysis will be carried out in the Review Manager 5.4 software. An adaptation of the GRADE framework for prognostic studies will be employed to judge the quality of evidence for each outcome reported in this systematic review [27].

## Discussion

This systematic review will summarize current evidence about STE-derived measures in the ESRD patients. Studies reported associations between STE-derived measures and all-cause and cardiovascular mortality and CV outcomes, but, is not fully understood the potential prognostic value of this diagnostic tool in ESRD patients. Evidence concerning measures (circumferential strain, radial strain) or 3D STE-derived indices will also be investigated. Therefore, this systematic review will also be displayed important knowledge gaps in the current literature regarding the possible utility of myocardial deformation indices in ESRD patients.

The mechanisms of decreased LV function in CKD patients are complex and not thoroughly understood [28] but CKD patients undergo a progressive change in myocardial composition and function due to pressure and volume overload, ischemia and CKD-related biochemical abnormalities.

The increased risk for cardiovascular morbidity and mortality might be caused by uremic cardiomyopathy (UC), a common complication in patients with ESRD that is characterized by cardiac fibrosis, capillary rarefaction, left ventricular hypertrophy, and both systolic and diastolic dysfunction [12-14, 29]. Therefore, early detection of UC by STE in patients with ESRD might identify patients at risk.

Ejection fraction (EF) measured from conventional echocardiogram is the gold standard for assessment of LV systolic function. However, EF has important technical limitations including load and geometry dependency and lacks sensitivity to detect subtle LV dysfunction [30].

Global longitudinal strain (GLS) is an emerging technique for measuring more subtle disturbances in LV systolic function. The two-dimensional strain method uses frame-by-frame tracking of naturally occurring acoustic reflections (speckles) that move with cardiac segments during the cardiac cycle; allowing measurement and characterization of regional and global systolic function. Less negative longitudinal strain (peak global) was associated with increased cardiovascular mortality. A more positive, and hence more abnormal GLS, was also associated with cardiac death [31-33].

Although radial, circumferential, and longitudinal fibers are predominantly in different layers of the myocardium, there is evidence of reduced myocardial deformation in all these orientations when we compared ESRD patient cohort to healthy persons [29]. As heart muscle is incompressible, strain parameters are interrelated in three dimensions, thus, by the time the chamber contracts and shortens in systole (circumferential and longitudinal strain), the wall thickens (radial strain) [34].

This systematic review will provide essential data regarding STE-derived measures as prognostic indicators of mortality and CV events in End-stage renal disease patients. The clinical implications of this work relate to the current status of diagnosis and treatment of heart disease in patients receiving dialysis.

### Availability of data and materials

The datasets generated and/or analyzed during the current study are available from the corresponding author on reasonable request.

## Supporting information

Supplementary_file_1_PRISMA-P_Checklist

Supplementary_file_2_Search_strategy

Supplementary_file_3_Eligibility_and_Data_Extraction_Forms

Supplementary_file_4_Quality_Assesment_Scale

## Data Availability

All data generated or analyzed during this study are included in this published article [and its supplementary information files].

## List of Abbreviations

AF: Atrial fibrillation
AICD: Automatic implantable cardioverter defibrillator
CHD: Coronary heart disease
CMR: Cardiovascular magnetic resonance imaging
CRS: Cardiorenal syndrome
CVD: Cardiovascular disease
DM: Diabetes mellitus
EF: Ejection fraction
ESRD: End-Stage Renal Disease
GLS: Global Longitudinal Strain
HD: Hemodialysis
HF: Heart failure
LILACS: Latin American and Caribbean Literature in Health Sciences
LV: Left ventricular
MI: Myocardial infarction
NOS: Newcastle-Ottawa Quality Assessment Scale of cohort studies
PRISMA-P: Preferred Reporting Items for Systematic Reviews and Meta-Analyses Protocols
PROSPERO: International registry of systematic reviews
RRT: Renal replacement therapy
STE: Speckle-tracking echocardiography
UC: Uremic cardiomyopathy

## Declarations

### Ethics approval and consent to participate

Not applicable.

### Consent for publication

Not applicable.

### Competing interests

The authors declare that they have no competing interests.

### Funding

None.

### Authors’ information

#### Affiliations

Clinical Analysis and Biomedicine Department, State University of Maringa, PR, BR.

JJVT.

Clinical Analysis Department, Federal University of Santa Catarina (UFSC), Florianópolis, SC, BR.

IGD.

Graduate Program in Biosciences and Pathophysiology (PBF-UEM), State University of Maringa, PR, BR.

BLSGM, JJVT, RPTO.

AGSM, EPB, GCD, JVD, MADS, RTPO.

Medicine Department, State University of Maringa, PR, BR.

RTPO, SSY.

Research Group on Systemic Arterial Hypertension, Arterial Stiffness and Vascular Aging (GPHARV-UEM), Regional University Hospital, State University of Maringa, PR, BR.

BLSGM, RTPO, SSY.

Uningá, Inga University Center, Maringa, PR, BR. JVD.

### Corresponding author

Correspondence to Barbara de Moura.

### Authors’ contributions

BLSGM has prepared this manuscript with clinical and methodological support from RTPO and JJVT, respectively. This manuscript has been reviewed by AGSM, EPB, GCD, JVD, MADS, IGD, SSY, RTPO and JJVT and edited by IGD, SSY, RTPO and JJVT. BLSGM is the corresponding author and has registered the protocol with the PROSPERO database. All authors agreed to be accountable for all aspects of this work and approved the final submission.

### Supplementary information

Supplementary file 1.

PRISMA-P Checklist.

Supplementary file 2.

Draft Search strategy.

Supplementary file 3.

Eligibility Form / Data Extraction Form.

Supplementary file 4.

Modified NOS Risk of Bias assessment.

## Notes

### Competing Interest Statement

The authors have declared no competing interest.

### Clinical Protocols

https://www.crd.york.ac.uk/prospero/display_record.php?ID=CRD42020222505

### Funding Statement

This study did not receive any funding

