## Supplementary_file_2_Search_strategy for "Prognostic implications of Global Longitudinal Strain versus Ejection Fraction in End-stage Renal Disease: a systematic review protocol"

Draft search strategy to be used for the PubMed online database

| **Search query:** Is the Global Longitudinal Strain a better prognostic marker than Ejection Fraction in the End Stage Renal Disease? | | | |
| --- | --- | --- | --- |
| **Sources searched: PubMed** | | | |
| **Limits: None**   - Date: 23/12/2020   Search number, Query, Sort By, Search details, Results | | | |
| Block 1 | 1 | "global longitudinal strain""[All Fields]" | 2330 |
|  | 2 | "speckle tracking echocardiography""[All Fields]" | 2994 |
| Block 2 | 3 | "chronic kidney disease""[All Fields]" | 53178 |
|  | 4 | ""haemodialysis""[All Fields] OR ""renal dialysis""[MeSH Terms] OR (""renal""[All Fields] AND ""dialysis""[All Fields]) OR ""renal dialysis""[All Fields] OR ""hemodialysis""[All Fields]" | 157888 |
|  | 5 | “end stage renal disease""[All Fields]" | 34005 |
| 6  (1 OR 2) | | "(""global longitudinal strain"") OR (""speckle tracking echocardiography"")",,,"""global longitudinal strain""[All Fields] OR ""speckle tracking echocardiography""[All Fields]" | 4549 |
| 7  (3 OR 4 OR 5) | | "((""chronic kidney disease"") OR (hemodialysis)) OR (""end stage renal disease"")",,,"""chronic kidney disease""[All Fields] OR (""haemodialysis""[All Fields] OR ""renal dialysis""[MeSH Terms] OR (""renal""[All Fields] AND ""dialysis""[All Fields]) OR ""renal dialysis""[All Fields] OR ""hemodialysis""[All Fields]) OR ""end stage renal disease""[All Fields]" | 210350 |
| 8  (6 AND 7) | | "((""global longitudinal strain"") OR (""speckle tracking echocardiography"")) AND (((""chronic kidney disease"") OR (hemodialysis)) OR (""end stage renal disease""))",,,"(""global longitudinal strain""[All Fields] OR ""speckle tracking echocardiography""[All Fields]) AND (""chronic kidney disease""[All Fields] OR (""haemodialysis""[All Fields] OR ""renal dialysis""[MeSH Terms] OR (""renal""[All Fields] AND ""dialysis""[All Fields]) OR ""renal dialysis""[All Fields] OR ""hemodialysis""[All Fields]) OR ""end stage renal disease""[All Fields])" | 122 |
