## Supplementary_file_4_Quality_Assesment_Scale for "Prognostic implications of Global Longitudinal Strain versus Ejection Fraction in End-stage Renal Disease: a systematic review protocol"

**NEWCASTLE - OTTAWA QUALITY ASSESSMENT SCALE**

**COHORT STUDIES (Modified)**

Note: A study can be awarded a maximum of one star for each numbered item within the Selection and Outcome categories.

The total quality score will be calculated by averaging the score of the two researchers with the range from 0 (lowest quality) to 6 (highest quality) and will be provided as a descriptive variable of the included studies.

**Selection**

1) Representativeness of the cohort

a) truly representative of the average person in the community **🟑**

b) somewhat representative of the average person in the community **🟑**

c) selected group of users eg nurses, volunteers

d) no description of the derivation of the cohort

2) Completeness of collection of potential confounders* (i.e. CVD risk factors)

*Based on Framingham risk factors modified to include ethnicity: age, sex, blood pressure, antihypertensive medication (yes/no), total cholesterol, HDL cholesterol (or BMI), smoking, diabetes mellitus, and ethnicity.

a) complete **🟑**

b) partially complete (≥5)**🟑**

c) partially complete (≤5)

d) no description

3) Demonstration that outcome of interest was not present at start of study (and/or sensitively analysis)

a) yes **🟑**

b) no

**Outcome**

4) Assessment of outcome

a) independent blind assessment **🟑**

b) record linkage **🟑**

c) self report

d) no description

5) Was follow-up long enough for outcomes to occur

a) yes (an adequate follow up period for outcome of interest is a year) **🟑**

b) no

6) Adequacy of follow up of cohorts

a) complete follow up - all subjects accounted for **🟑**

b) subjects lost to follow up unlikely to introduce bias - small number lost - > 80% follow up, or description provided of those lost **🟑**

c) follow up rate < 80% and no description of those lost

d) no statement

**NEWCASTLE - OTTAWA QUALITY ASSESSMENT SCALE COHORT STUDIES (Modified)**

| **First Author, Year** | **SELECTION** | | | **OUTCOME** | | | **TOTAL QUALITY SCORE** |
| --- | --- | --- | --- | --- | --- | --- | --- |
|  | **Representativeness of the cohort** | **Completeness of collection of potential confounders*** | **Demonstration that outcome of interest was not present at start of study (and/or sensitivity analysis)** | **Assessment of outcome** | **Was follow-up long enough for outcomes to occur** | **Adequacy of follow up of cohorts** | **0 - 6** |
|  | a) truly representative of the average person in the community 1  b) somewhat representative of the average person in the community 1  c) selected group of users 0  d) no description of the derivation of the cohort 0 | a) complete 1  b) partially complete (>5)1  c) partially complete (≤5) 0  d) no description 0 | a) yes 1  b) no 0 | a) independent blind assessment 1  b) record linkage 1  c) self-report 0  d) no description 0 | a) yes** 1  b) no 0 | a) complete follow up - all subjects accounted for 1  b) subjects lost to follow up unlikely to introduce bias - small number lost - > 80% follow up, or description provided of those lost 1  c) follow up rate < 80% and no description of those lost 0  d) no statement 0 | lowest quality to highest quality |

From: Wells G, Shea B, O’Connell D, Peterson J, Welch V, Losos M, Tugwell P. Newcastle-Ottawa quality assessment scale cohort studies. Available on: http://www. ohri. ca/programs/clinical_epidemiology/oxford. Asp.

*Based on Framingham risk factors modified: age, sex, blood pressure, antihypertensive medication (yes/no), total cholesterol, HDL cholesterol (or BMI), smoking, diabetes mellitus, and ethnicity.

**An adequate follow up period for outcome of interest is a year.
